# Repeat controlled human malaria infection of healthy UK adults with blood-stage *Plasmodium falciparum*: safety and parasite growth dynamics

**DOI:** 10.1101/2022.06.27.22276860

**Authors:** Jo Salkeld, Yrene Themistocleous, Jordan R. Barrett, Celia H. Mitton, Thomas A. Rawlinson, Ruth O. Payne, Mimi M. Hou, Baktash Khozoee, Nick J. Edwards, Carolyn M. Nielsen, Diana Muñoz Sandoval, Florian A. Bach, Wiebke Nahrendorf, Raquel Lopez Ramon, Megan Baker, Fernando Ramos-Lopez, Pedro M. Folegatti, Doris Quinkert, Katherine J. Ellis, Ian D. Poulton, Alison M. Lawrie, Jee-Sun Cho, Fay L. Nugent, Philip J. Spence, Sarah E. Silk, Simon J. Draper, Angela M. Minassian

## Abstract

In endemic settings it is known that natural malaria immunity is gradually acquired following repeated exposures. Here we sought to assess whether similar acquisition of blood-stage malaria immunity would occur following repeated parasite exposure by controlled human malaria infection (CHMI). We report the findings of a repeat homologous blood-stage *Plasmodium falciparum* (3D7 clone) CHMI study (VAC063C; ClinicalTrials.gov NCT03906474). In total, 24 healthy, unvaccinated, malaria-naïve UK adult participants underwent primary CHMI followed by drug treatment. Ten of these then underwent secondary CHMI in the same manner, and then six of these underwent a final tertiary CHMI. As with primary CHMI, malaria symptoms were common following secondary and tertiary infection, however, most resolved within a few days of treatment and there were no long term sequelae or serious adverse events related to CHMI. Despite detectable induction and boosting of anti-merozoite serum IgG antibody responses following each round of CHMI, there was no clear evidence of anti-parasitic immunity (manifest as reduced parasite growth *in vivo*) conferred by repeated challenge with the homologous parasite in the majority of volunteers. However, three volunteers showed some variation in parasite growth dynamics *in vivo* following repeat CHMI that were either modest or short-lived. We also observed no major differences in clinical symptoms or laboratory markers of infection across the primary, secondary and tertiary challenges. However, there was a trend to more severe pyrexia after primary CHMI and the absence of a detectable transaminitis post-treatment following secondary and tertiary infection. We hypothesize that this could represent the initial induction of disease tolerance or clinical malaria immunity. Repeat homologous blood-stage CHMI is thus safe and provides a model with the potential to further the understanding of the acquisition of blood-stage immunity in a highly controlled setting.

## Introduction

Controlled human malaria infection (CHMI) is the most developed experimental human microbial infection model. It has been critical to advancing new therapeutic drugs and vaccines, including the most advanced anti-sporozoite vaccine for *Plasmodium falciparum*, RTS,S/AS01, and has provided key insights into parasite immuno-biology and pathogenesis of disease (1). CHMI was pioneered in the fascinating era of malariotherapy, in which deliberate malaria infection, delivered either via mosquito bite or by blood transfusion, was administered as treatment for neurosyphilis prior to the availability of penicillin in the mid-1940s (2). Since it was the induction of fever that led to improvement of neurosyphilitic symptoms, patients were routinely re-challenged with both homologous and heterologous strains (with *P. vivax* and to a lesser extent with *P. falciparum*) and this became the standard treatment of neurosyphilis. Retrospective examinations of reinfection data in patients using homologous *P. vivax* or homologous and/or heterologous *P. falciparum* demonstrated a reduction in fever episodes (“clinical immunity”) during the secondary infection as well as reductions in parasitaemia (“anti-parasitic immunity”), suggesting some partial immunity can be conferred from one previous parasite exposure (3, 4). However, this phenomenon has not been re-explored by CHMI in the “modern era”.

In the natural exposure setting (i.e. in malaria-endemic countries) the manifestations of severe malaria disease vary with age and transmission setting (5). Nonetheless, immunity to severe malaria is gradually acquired by young children following repeated infections with *P. falciparum*, and ultimately older children and adults typically develop asymptomatic infections whereby they are able to tolerate much higher levels of parasitaemia without symptoms or clinical signs of malaria (6). This naturally-acquired immunity, observed in older children and adults, is widely understood to involve a complex interplay of cellular and humoral immune mechanisms, and to develop over time, following repeated exposures, to multiple parasite strains.

Repeat homologous blood-stage CHMI (with malaria of any species) has the potential to further the understanding of the impact of prior malaria exposure on a subsequent infection in a highly controlled and experimental setting. However, prior to recent studies in Oxford, only one previous study had administered a second intravenous *P. falciparum* blood-stage challenge in humans (7). Five malaria-naïve volunteers were repeatedly inoculated with infected erythrocytes at low doses, followed by administration of anti-malarial drugs before development of clinical infection. Three of the four volunteers who completed the study were protected from infection after three rounds of low dose blood-stage challenge and cure, with no parasite DNA detected by quantitative PCR (qPCR) after the fourth inoculation. However, with very small numbers, lack of a control group and detection of residual atovaquone, which may have confounded the observed outcome, clear interpretation of these results is difficult (7, 8). In contrast, many healthy adult volunteers, in other malaria vaccine clinical trials worldwide, have been re-challenged with *P. falciparum* malaria delivered by mosquito bite (9-12) or as injected cryopreserved sporozoites (13-15). However, in most cases these were vaccinees who had shown evidence of vaccine-induced sterilizing or partial immunity following their primary CHMI. These studies also aimed to assess the durability of vaccine-induced pre-erythrocytic immunity as opposed to any impact on the subsequent blood-stage of infection.

More recently, as part of the VAC063 RH5.1/AS01_B_ blood-stage vaccine efficacy trial (16), we administered a secondary homologous *P. falciparum* (3D7 clone) blood-stage challenge to a subset of unvaccinated infectivity control volunteers with unexpected results. Upon primary CHMI in the part of the study called “VAC063A”, all fifteen infectivity controls were diagnosed at ∼10,000 parasites per mL of blood within 8-12 days as routinely seen in this CHMI model in malaria-naïve adults. Subsequently, following secondary homologous CHMI approximately 4 months later (called “VAC063B”), six out of eight volunteers showed identical blood-stage parasitaemia to the primary infection, however, two out of eight participants demonstrated a reduction in their parasite multiplication rate (PMR) as compared to primary infection and the remainder of the secondary CHMI cohort. Indeed, one of these two volunteers showed undetectable blood-stage infection out to 19 days.

Given natural immunity to the blood-stage parasite is slow to acquire, these findings after a single acute primary exposure were not anticipated. However, they did suggest that repeat homologous blood-stage CHMI could provide a model to further interrogate the immunological mechanisms that underlie the acquisition of clinical and anti-parasitic immunity, which could then guide development of new vaccination strategies. Here, we therefore sought to further develop this model by inviting these previous participants back to take part in a final follow-on challenge study (called “VAC063C”), to enable the assessment of safety and parasite growth dynamics following tertiary, secondary and primary homologous blood-stage CHMI.

## Results

### Participant flow and demographics

Twenty-five volunteers were screened for the VAC063C study. Sixteen of these were malaria-naïve healthy adult volunteers, and the other nine volunteers were screened following invitation to participate, after their previous enrolment and receipt of blood-stage *P. falciparum* CHMI as unvaccinated infectivity controls in the VAC063 study (A and B CHMIs, ClinicalTrials.gov NCT02927145) (16) (**Figure 1A**).

**Figure 1.**
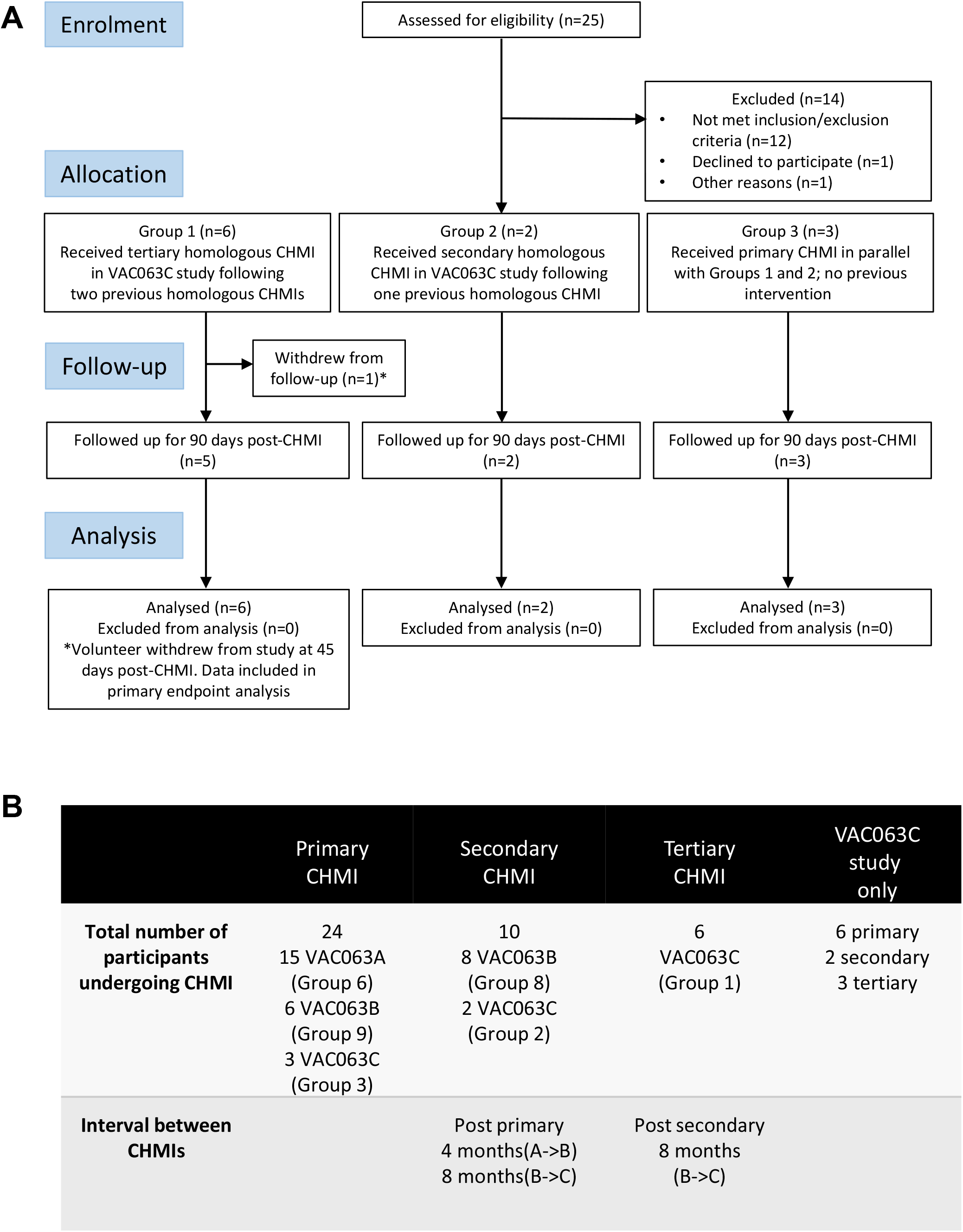
VAC063C flow chart of study design and volunteer recruitment. **(A)** Enrolment into the VAC063C study began in October 2018. Twenty five volunteers were screened for eligibility. Eleven eligible volunteers were identified, six of whom were VAC063 participants returning for a tertiary controlled human malaria infection (CHMI) (Group 1), two of whom were VAC063 participants returning for a secondary CHMI (Group 2) and three of whom were newly enrolled participants for primary CHMI (Group 3). Clinical follow-up continued until 90 days after challenge (C+90) and was completed by 14^th^ January 2019. Volunteer demographics are summarized in **Table S1. (B)** In total (across three CHMI studies, VAC063A, B and C) 24 participants underwent primary CHMI, 10 of whom subsequently underwent secondary CHMI and six of those participants returned for a tertiary CHMI. The first CHMI (VAC063A) was on 14^th^ November 2017, the second (VAC063B) was on 6^th^ March 2018 and the third (VAC063C) was on 6^th^ November 2018.

Eleven volunteers were subsequently enrolled, comprising six volunteers who had previously undergone two CHMIs in the VAC063 study (Group 1), two volunteers who had received a single prior CHMI (Group 2) and a further three malaria-naïve volunteers to act as primary infectivity controls (Group 3). Over the entire study period, only a single participant withdrew prior to completion (after 45 days of follow-up post-CHMI) due to personal reasons. **Table S1** compares the demographics of those undergoing primary, secondary or tertiary CHMI in the VAC063C study as well as the pooled demographic data across all three CHMI studies (VAC063A, B and C). Age, gender, ethnicity and body mass index (BMI) were comparable across all participants undergoing primary, secondary and tertiary CHMI.

### Period of controlled human malaria infection

Blood-stage CHMI, with the *P. falciparum* 3D7 clone, was initiated for the VAC063C study on 6^th^ November 2018; this was approximately 8 months after completion of the preceding VAC063B CHMI (**Figure 1B**). Exactly as for VAC063A and VAC063B, the CHMI was initiated by administration of the inoculum to each participant via an intravenous injection. This contained approximately 777 parasitised erythrocytes in 5 mL of normal saline, as estimated by a limiting dilution assay of the inoculum (executed as soon as the final participant had received it). This was highly comparable to the dose of approximately 452 and 857 parasitised erythrocytes administered to participants during the VAC063A and B CHMIs, respectively (16). All eleven participants subsequently developed patent blood-stage parasitaemia and were diagnosed and treated at pre-defined parasitaemia/clinical thresholds. All participants completed the course of antimalarials as prescribed and all follow-up visits were completed by 14^th^ February 2019.

### Safety of repeat homologous *P. falciparum* CHMI

There were no serious adverse events (SAEs) or unexpected reactions deemed possibly, probably or definitely related to CHMI, blood draws or study drugs during the course of the VAC063C study and no participants withdrew due to study-related adverse events (AEs). Otherwise, only a single SAE deemed unrelated to study interventions occurred during VAC063C; abdominal pain secondary to suspected renal calculus requiring overnight hospital admission. We next proceeded to analyse the safety data from the VAC063C study alone, as well as data from all participants combined across the three VAC063 blood-stage CHMI trials. In total 24 participants underwent one CHMI only, 10 volunteers received two homologous CHMIs and 6 received three homologous CHMIs. The intervals between the CHMI periods were approximately 4 months (VAC063A to B) and 8 months (VAC063B to C) (**Figure 1B**).

The maximum severity reported and relative frequencies of CHMI-related solicited AEs for all participants receiving primary, secondary and tertiary CHMI across the three CHMI studies are shown in **Figure 2A,C**, with unsolicited AEs shown in **Table S2**. Data on solicited AEs from the VAC063C study only are shown in **Figure 2B**. There were no major differences in the number or type of AE reported across the primary, secondary and tertiary CHMI groups, although severe AEs were limited to those undergoing primary or secondary CHMI (with the caveat that a smaller number of participants underwent tertiary CHMI). The most commonly reported AEs were headache, fatigue, malaise, followed by feverishness, chills, sweats, myalgia, nausea and arthralgia. A minority of volunteers reported diarrhoea or vomiting. Maximum severity of AEs peaked between 24 and 48 hours post-diagnosis (after starting antimalarial drug treatment) and most AEs had resolved within a few days of CHMI or completing treatment with no long-term sequelae (**Figure 3**). A minority of participants also reported AEs possibly related to taking antimalarial treatment (artemether/lumefantrine or atavoquone/proguanil). These occurred in the 24-48 hour period after initiating treatment and resolved quickly (**Figure S1**).

**Figure 2.**
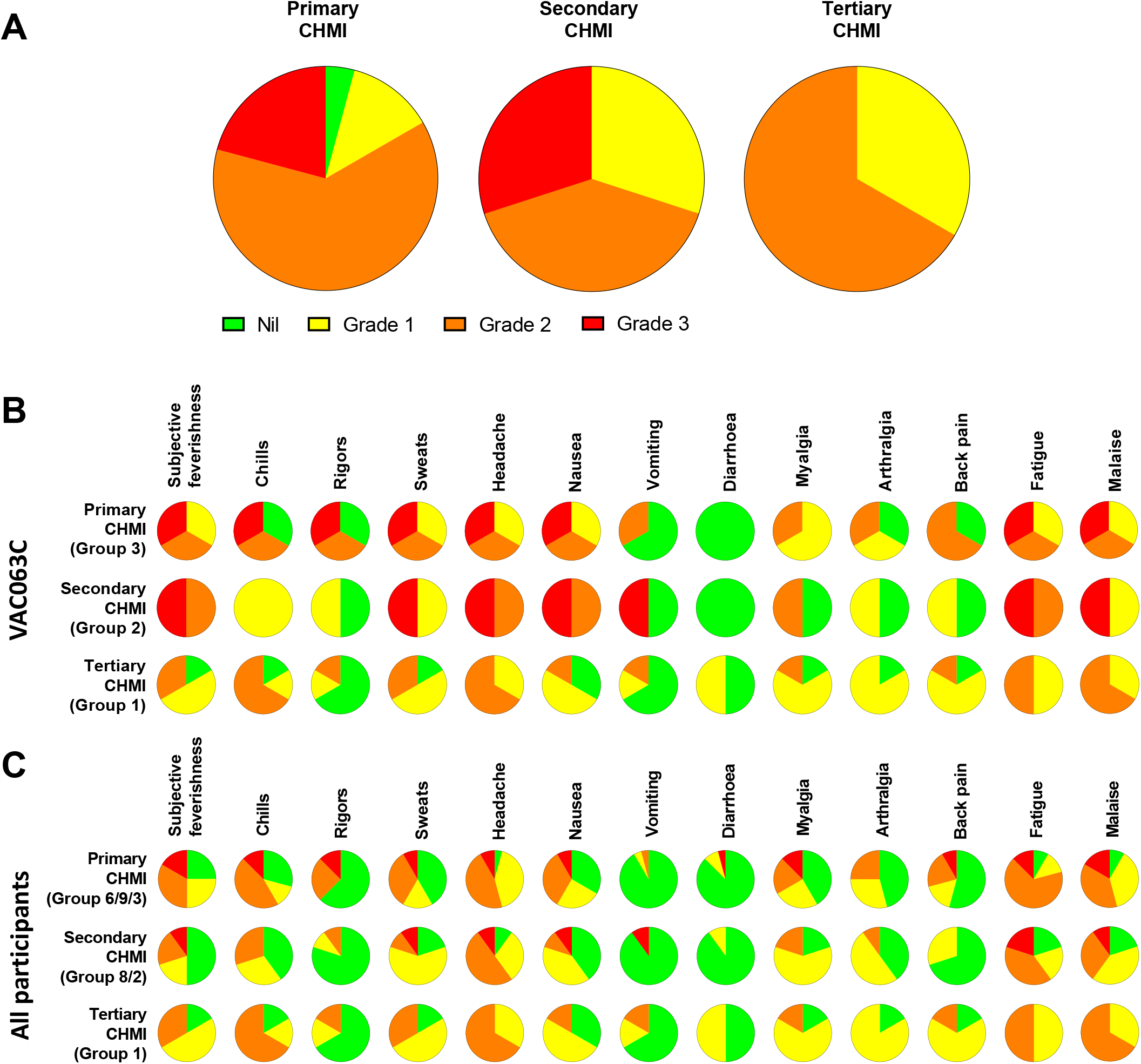
Safety of repeated *P. falciparum* blood-stage CHMI. Participants were asked about the presence of 13 solicited systemic adverse events (AE) at study each visit following CHMI. (**A**) The maximum severity of any solicited AE reported by each participant in the 48 hours before and after diagnosis during primary (n=24), secondary (n=10) and tertiary (n=6) CHMI is shown as a proportion of the total number of participants. (**B**) VAC063C: The solicited AEs recorded during the CHMI period are shown as the maximum severity reported by each participant and as a proportion of the participants reporting each individual AE for primary (n=3), secondary (n=2) and tertiary (n=6) CHMI. AE data was collected until 90 days after challenge. Colour coding as per panel A. (**C**) The solicited AEs recorded during the CHMI period are shown as the maximum severity reported by each participant and as a proportion of the participants reporting each individual AE for primary (n=24), secondary (n=10) and tertiary (n=6) CHMI for all participants across the three CHMI studies. Groups refer to specific study group numbers in VAC063A, B and C. Colour coding as per panel A.

**Figure 3.**
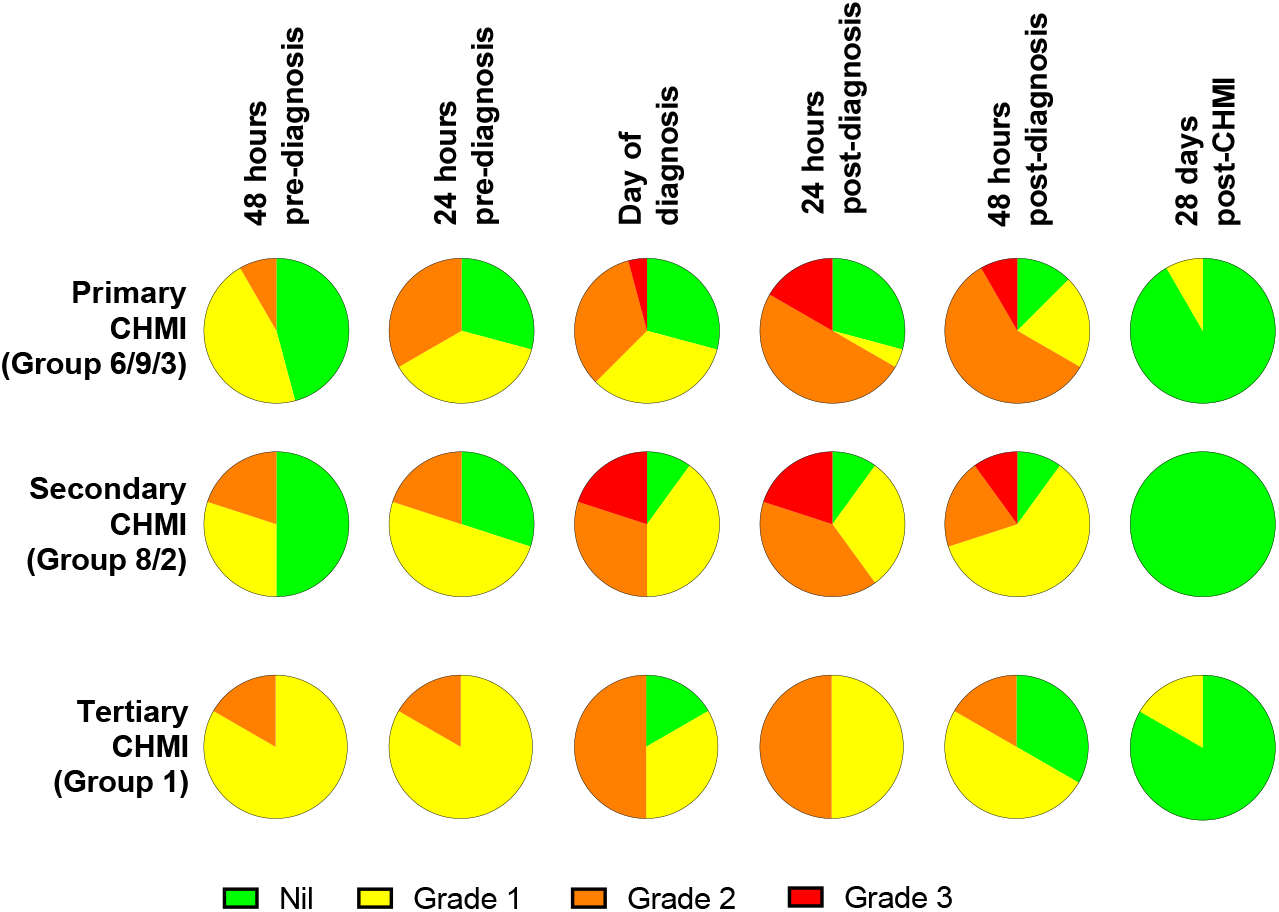
Symptom evolution during repeated *P. falciparum* blood-stage CHMI. The maximum severity of any solicited systemic adverse events (AE) recorded at the indicated time points during the CHMI period for each participant for primary (n=24), secondary (n=10) and tertiary (n=6) CHMI.

We next analysed the frequency and severity of objective clinical observations and laboratory AEs (**Figure 4, Table S3**). We observed no differences in haemoglobin or platelet counts across the three cohorts undergoing repeat *P. falciparum* infection. However, lymphocytopaenia was more frequent following secondary and tertiary CHMI (affecting >60% participants) compared to <30% following primary CHMI. There was also a trend toward less severe pyrexia with each successive CHMI (**Figure 4A,C**). In nearly all cases, these clinical changes were transient, coinciding with malaria diagnosis, with pyrexia resolving within the next 48 hours and lymphocytes normalizing by 6 days post-treatment (T+6) in VAC063C or by 28 days post-CHMI (C+28) in VAC063A and B (noting T+6 was not assessed in these earlier studies).

**Figure 4.**
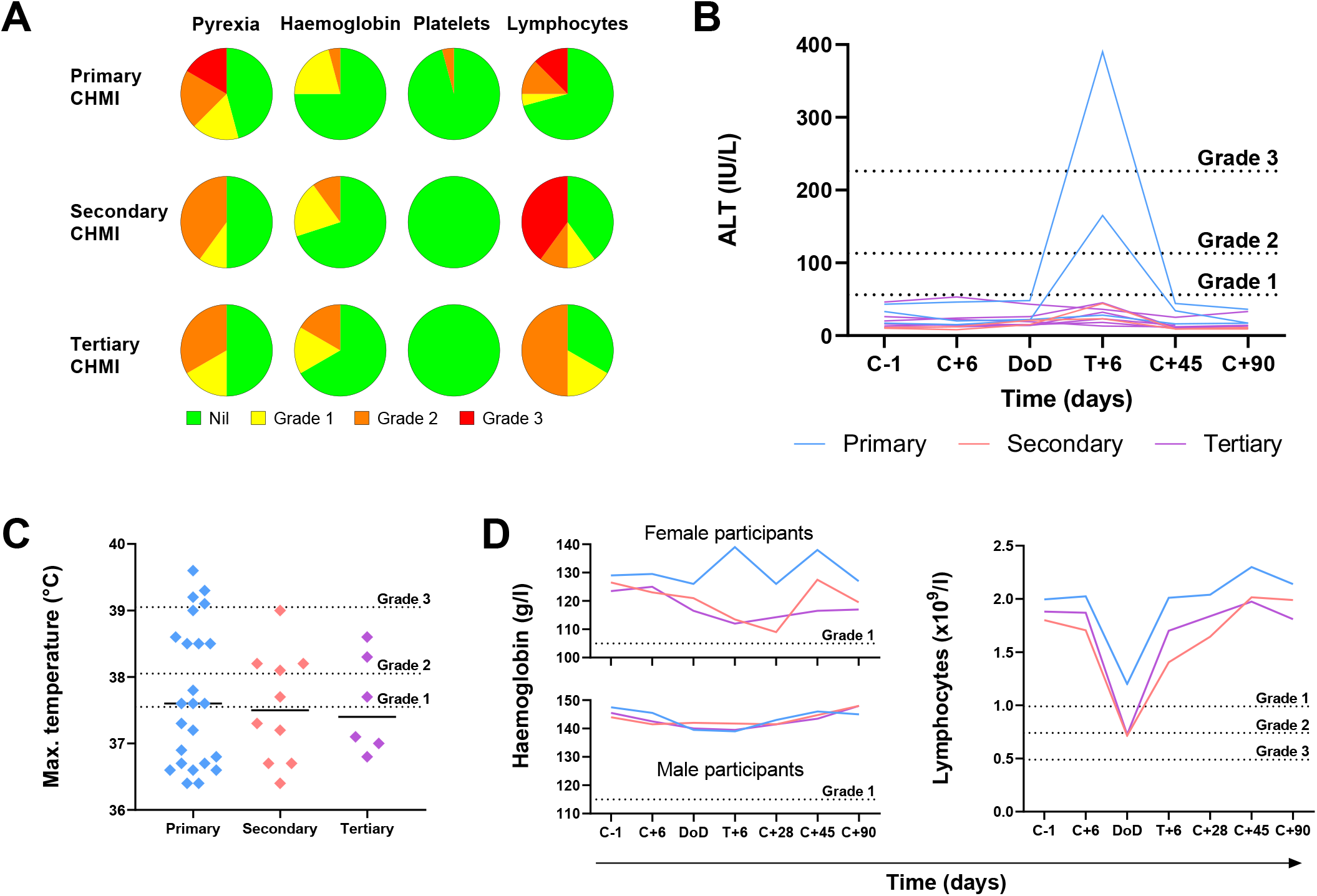
Objective measurements and laboratory AEs during repeated *P. falciparum* blood-stage CHMI. (**A**) Frequency and severity of pyrexia, anaemia, thrombocytopaenia and lymphopaenia during primary (n=24), secondary (n=10) and tertiary (n=6) CHMI from screening until 90 days post-CHMI. (**B**) Alanine aminotransferase (ALT) for each participant in VAC063C at each timepoint for primary (n=3), secondary (n=2) and tertiary (n=6) CHMI showing significant transaminitis after treatment of the primary infection (*P* = 0.03 as calculated by Kruskal-Wallis test of peak ALT values). Dashed lines show the local cut off for Grade 1, 2 and 3 abnormalities. Time is number of days post-CHMI, except for day of diagnosis (DoD) and day 6 post-treatment (T+6) which vary by participant. The same information for VAC063 A and B (which did not include a T+6 assessment) is presented in **Figure S2**. (**C**) Maximum recorded temperature during primary (n=24), secondary (n=10) and tertiary (n=6) CHMI. Individual data points and the median are shown. Dashed lines show the local cut off for Grade 1, 2 and 3 abnormalities. Kruskal-Wallis test showed no significant difference between primary, secondary and tertiary CHMI (*P* = 0.31). (**D**) Median haemoglobin concentration and lymphocyte count for all participants across VAC063A, B and C over time for primary (n=24), secondary (n=10) and tertiary (n=6) CHMI (of which n=18 were female and n=22 male). Time points are the same as in panel B. Kruskal-Wallis test showed no significant difference between primary, secondary and tertiary CHMI (minimum haemoglobin concentration *P* = 0.93; minimum lymphocyte count *P* = 0.19).

During VAC063C we also noted a transaminitis occurring in Group 3 participants (those undergoing primary CHMI), as measured by significantly raised alanine aminotransferase (ALT), one severe, at 6 days post-diagnosis and treatment (T+6) (**Figure 4B**); *P* = 0.03 by Kruskal-Wallis test. This was not apparent in any participant undergoing secondary or tertiary CHMI. We potentially failed to observe this degree of transamintis in the VAC063A and VAC063B cohorts because these studies did not include the T+6 assessment (**Figure S2**), although four milder elevations of ALT were observed. Further examination of the two volunteers with transaminitis in VAC063C revealed similar increases in aspartate aminotransferase (AST) and gamma-glutamyl transferase (GGT), and that the participants had neither signs nor symptoms of hepatitis. One participant reported mild loss of appetite and, upon examination, mild epigastric and suprapubic tenderness were present but without right upper quadrant pain or hepatosplenomegaly. Their AST normalised after 3 days and ALT after 12 days, although GGT did not fully normalise until the last study visit (day C+90). The second participant remained completely asymptomatic and all transaminases (ALT, AST and GGT) normalised after 28 days. In both participants bilirubin and markers of synthetic liver function (albumin, alkaline phosphatase (ALP) and coagulation screen) remained normal. Both participants had been EBV and hepatitis A virus IgG positive prior to CHMI, however, in order to rule out new blood-borne infections as the cause of liver derangement, both participants were screened for CMV, hepatitis B, hepatitis C, human immunodeficiency virus (HIV) and hepatitis E infection; all serology was negative.

### Parasite growth dynamics following repeat homologous *P. falciparum* CHMI

Across the three VAC063 blood-stage CHMI studies, all participants developed detectable blood-stage parasitaemia and were diagnosed according to clinical/parasitological criteria defined in the trial protocols. Individual parasitaemias were measured over time by qPCR, and are shown for all volunteers undergoing primary (n=24), secondary (n=10) and tertiary (n=6) CHMI (**Figure 5A, Table S4**). Analysis of the median qPCR result showed highly comparable acute blood-stage parasite growth across each repeat infection cohort (**Figure 5B**). We confirmed that volunteers were clinically diagnosed at similar levels of parasitaemia across each cohort (approximately 10,000 parasites per mL [p/mL] blood) with no significant differences observed; *P* = 0.79, Kruskal-Wallis test (**Figure 5C**).

**Figure 5.**
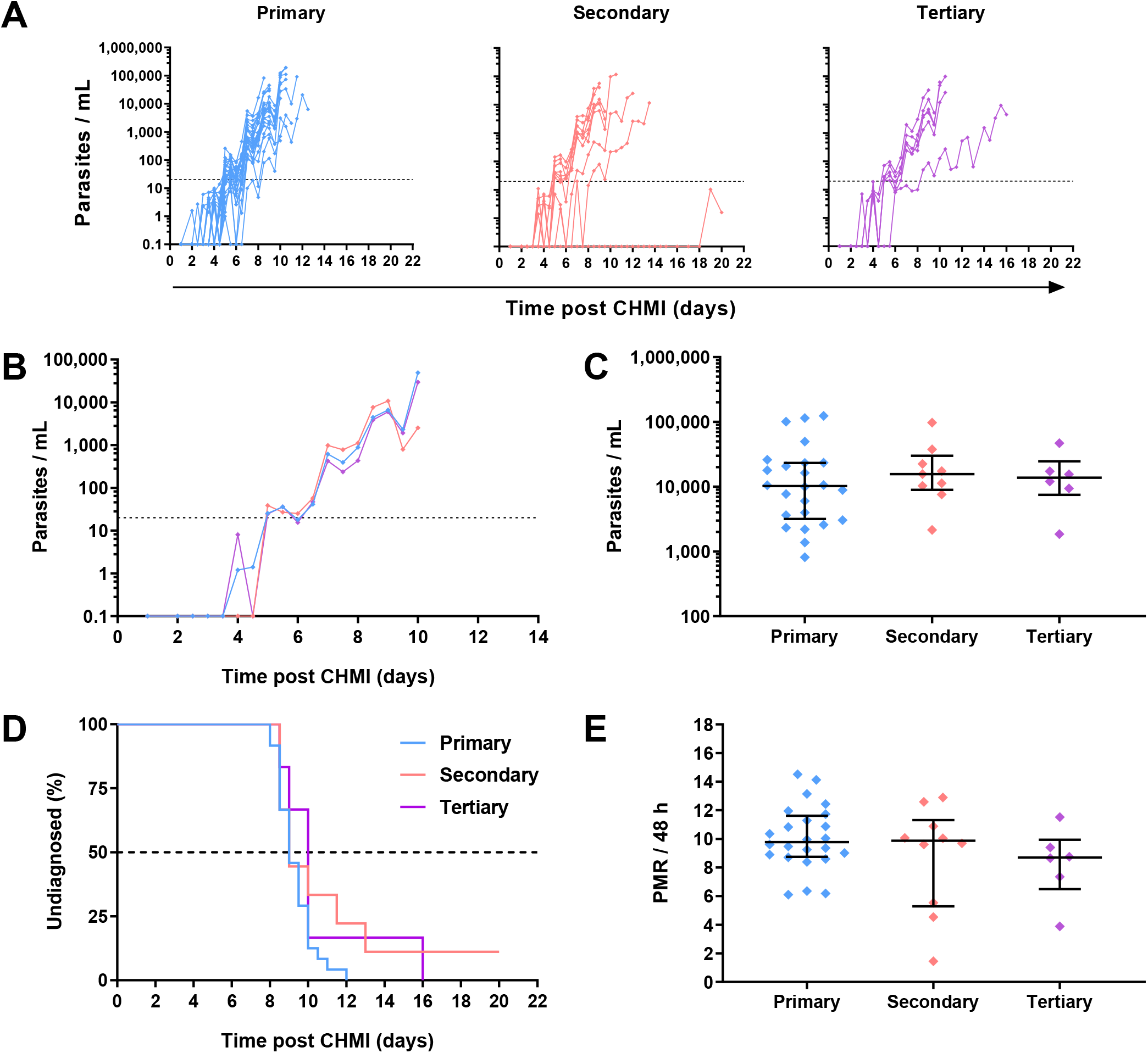
Parasite growth dynamics following repeated *P. falciparum* blood-stage CHMI. (**A**) qPCR data for primary (n=24), secondary (n=10) and tertiary (n=6) CHMI across the VAC063A, B and C studies. Parasitaemia measured by qPCR over time in parasites/mL (p/mL) blood is shown for each volunteer. CHMI was initiated by intravenous inoculation of *P. falciparum* infected erythrocytes on day 0. The lower limit of quantification is indicated by the dotted line at 20 p/mL; values below this level are plotted for information only. (**B**) Median qPCR data are shown for primary, secondary and tertiary CHMI. Colours as in panel A. (**C**) Parasitaemia at the time of diagnosis. Individual data points and the median ± inter-quartile range for primary (n=24), secondary (n=9) and tertiary (n=6) CHMI are shown. Note one participant requested antimalarial treatment at day C+20 during secondary CHMI without reaching the diagnostic criteria so no data point is shown for this participant. *P* value as calculated by Kruskal-Wallis test showed no significant difference between groups. (**D**) Kaplan-Meier plot of time to diagnosis in days. Note one secondary CHMI participant requested treatment and was censored at day C+20. Log-rank (Mantel-Cox) test showed no significant difference in time to diagnosis between primary, secondary and tertiary CHMI. (**E**) The parasite multiplication rate (PMR) per 48 hours was modelled from the qPCR data up until the time point of diagnosis. Individual data points and median ± inter-quartile range for primary, secondary and tertiary CHMI PMR are shown. *P* value as calculated by Kruskal-Wallis test showed no significant difference.

At the group level there was no evidence that prior blood-stage *P. falciparum* infection affected subsequent homologous parasite growth, as evidenced by comparable time-to-diagnosis (**Figure 5D**) across the three cohorts; *P* =0.16, log-rank test. The median time-to-diagnosis was 9 days post-primary CHMI (n =24), 9 days post-secondary CHMI (n=10), and 10 days post-tertiary CHMI (n=6). Similar results were observed for the VAC063C protocol pre-specified primary analysis of parasite growth dynamics by comparison of the parasite multiplication rate (PMR). Overall, there was no significant difference in the PMRs between the three groups. The median PMR per 48 h for primary CHMI was 9.8, 9.9 for secondary and 8.7 for tertiary, *P* = 0.3, Kruskal-Wallis test (**Figure 5E**).

Nevertheless, in contrast to the majority (7/10) of volunteers undergoing repeat CHMI who showed near identical blood-stage parasitaemia across all three infections (**Figure 6A**), we noted a small subset of three volunteers where some change in the phenotype of parasite growth occurred (**Figure 6B-D**). Two participants demonstrated slower blood-stage parasite growth following successive CHMIs. The first participant had a relatively low PMR following primary CHMI and then a consistent but modest reduction in PMR during subsequent parasite exposures (4-month then 8-month interval) with PMR per 48 h falling from 6.1 to 4.5 and then to 3.9 after the secondary and tertiary CHMIs, respectively (**Figure 6B**). The second volunteer had a 1.7-fold drop in their PMR per 48 h (9.4 to 5.5) between primary and secondary CHMI (administered 8 months apart), but did not go on to receive a tertiary challenge (**Figure 6C**).

**Figure 6.**
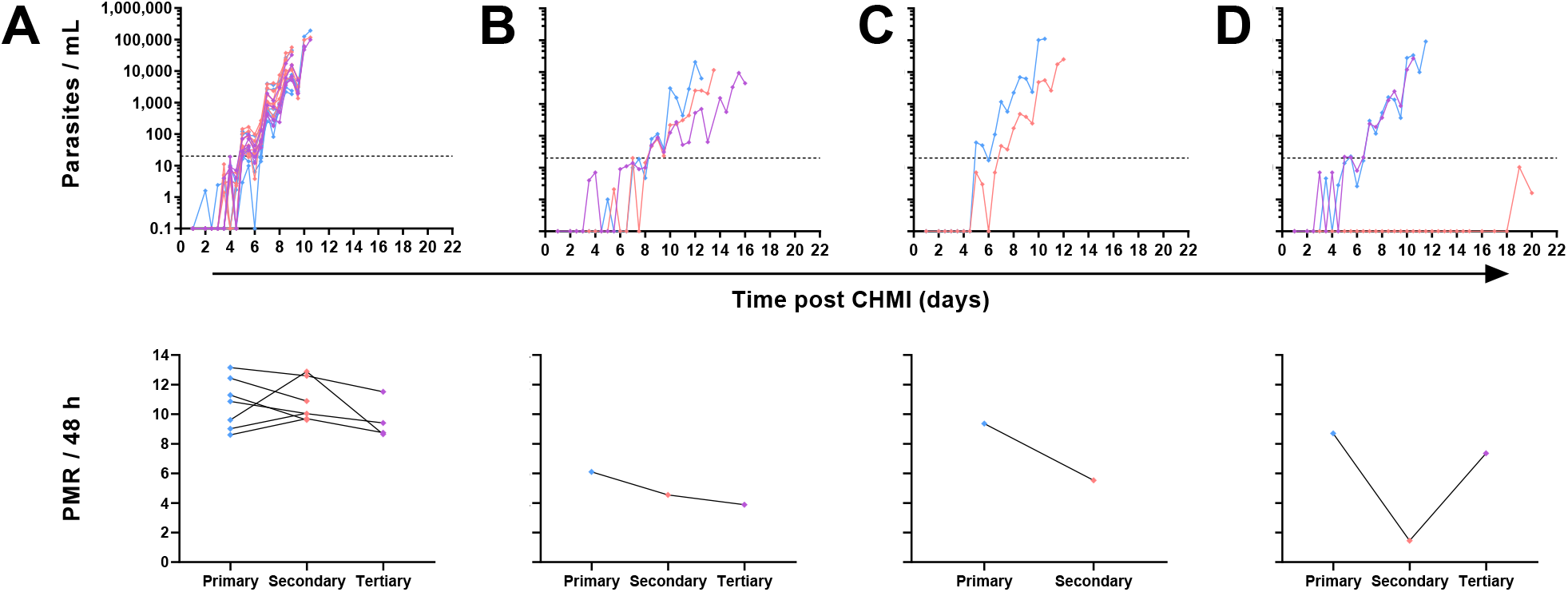
Individual variation in parasite growth dynamics of repeated *P. falciparum* blood-stage CHMI. qPCR data for primary (blue), secondary (pink) and tertiary (purple) CHMI. Parasitaemia is shown over time for each volunteer who underwent more than one CHMI (total n=10). The PMR per 48 hours was modelled from the qPCR data up until the time point of diagnosis for each participant for each CHMI and is shown below the relevant parasite growth graph. (**A**) Data from participants who showed minimal variation in parasite growth dynamics between each CHMI (n=7 for primary and secondary CHMI, of whom n=4 also underwent tertiary CHMI). (**B**) One participant showed consistently slower parasite growth with each subsequent CHMI. (**C**) One participant showed slower parasite growth during secondary CHMI compared to primary but did not undergo a tertiary CHMI. (**D**) One participant showed completely suppressed parasite growth on secondary CHMI until day C+19 but no change compared to primary upon tertiary CHMI.

Finally, one participant completely suppressed parasitaemia post-secondary CHMI (which occurred 4 months post-primary CHMI) until day C+19 (**Figure 6D**). At this point, blood parasitaemia was detected by qPCR for the first time but remained below the lower limit of quantification at 20 p/mL. A similar result was obtained on day C+20, however at this point the participant requested antimalarial treatment so was drug-treated at low-level parasitaemia and never met criteria for diagnosis. A pre-treatment blood sample was also cultured in the laboratory and outgrowth of *P. falciparum* parasites was subsequently confirmed *in vitro* (data not shown). However, upon tertiary CHMI during the VAC063C study (8 months later), this same volunteer developed blood-stage parasitaemia with essentially identical growth to that observed following their primary CHMI. Further clinical investigation yielded no obvious reason for suppressed parasite growth in the blood following this volunteer’s secondary CHMI, and a subsequent screen for the presence of antimalarial drugs in plasma proved negative. Indeed, all antimalarial compounds tested were below the limit of detection in all plasma samples tested, with the exception of lumefantrine. This gave a low-level signal across all samples tested, with almost identical within-individual results pre- and post-CHMI (samples taken 2-3 weeks apart). The significance of these very low levels of detectable lumefantrine in all samples remain unexplained but cannot explain the observed parasite growth rates given their presence in all samples.

### Antibody responses following repeat homologous CHMI

Finally, we assessed serum IgG antibody responses by ELISA to three commonly-studied merozoite antigens – the 19 kDa C-terminus of merozoite surface protein 1 (MSP1_19_), apical membrane antigen 1 (AMA1) and reticulocyte-binding protein homologue 5 (RH5). MSP1_19_-specific serum IgG responses were detectable at day C+28 in >90% volunteers following initial CHMI. These were subsequently boosted by each successive CHMI and were relatively well-maintained between parasite exposures (**Figure 7A**). In contrast, serum IgG responses to AMA1 were less dominant after the primary CHMI (with responses only detected in a minority of volunteers). These then boosted following repeat CHMI in a similar manner to MSP1_19_, however, these responses dropped back to baseline between each successive *P. falciparum* exposure (**Figure 7B**). Responses to RH5 were undetectable in all volunteers at all time-points tested (data not shown).

**Figure 7.**
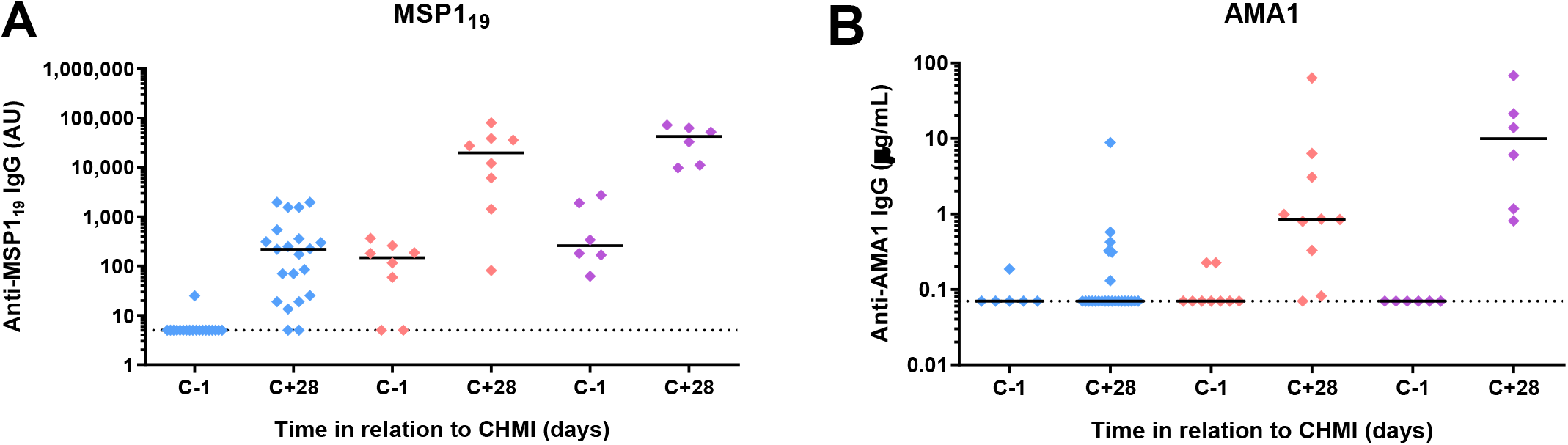
Induction of serum antibody responses to merozoite antigens during CHMI. (**A**) Serum anti-MSP1_19_ total IgG ELISA was conducted on samples from participants for primary (n=19 at dC-1; n=21 at dC+28), secondary (n=8) and tertiary (n=6) CHMI. Sera from the pre-CHMI (dC-1) and 28 days post-CHMI (C+28) time points were tested. **(B)** Serum anti-AMA1 total IgG as measured by standardized ELISA on samples from participants for primary (n=6 at dC-1; n=24 at dC+28), secondary (n=9 at dC-1; n=10 at dC+28) and tertiary (n=6) CHMI. Samples colour coded as per previous figures. Individual data points and median are shown.

We also screened serum samples from all participants undergoing more than one CHMI for the presence of red cell alloantibodies, which could have been raised by the repeated exposure to the blood-stage inoculum. Samples were tested both at baseline (before primary CHMI) and at final follow-up (day C+90) after the final (whether secondary or tertiary) CHMI. All tested samples were negative for alloantibodies (data not shown).

## Discussion

This is the first assessment of repeat homologous *P. falciparum* blood-stage CHMI in the modern era, where volunteers have reached diagnostic criteria before initiation of anti-malarial therapy. Overall we have demonstrated that repeated exposure to homologous blood-stage parasites by CHMI is safe in healthy adults, but does not lead to the development of any significant anti-parasitic blood-stage immunity in the vast majority of re-challengees.

The frequencies of CHMI-associated solicited AEs were within expected ranges in all participants undergoing a primary CHMI, consistent with previously reported studies of primary CHMI using both blood-stage and sporozoite infection models (16-19). Inference about relative frequency of solicited AEs in re-challenged participants is limited by relatively small sample size, selection bias of returning participants and inter-subject variability in reporting of AEs, however, our findings suggest that there are no major differences in the frequencies of AEs following homologous re-challenge, or any increased risk, as compared to primary CHMI. The majority of AEs also resolve within a few days of treatment.

There were also no statistically significant differences in the frequencies of objective clinical or laboratory markers of malaria infection across the repeat CHMIs, although we noted the highest levels of pyrexia occurred in a subset of volunteers undergoing primary CHMI. This is not inconsistent with the retrospective analyses of the old neurosyphilis treatment data (4), although the key difference here is that modern CHMIs only allow acute blood-stage infection prior to treatment in contrast to uncontrolled parasitaemia which almost certainly led to more effective acquisition of anti-malarial clinical immunity. We also noted the lowest lymphocyte counts occurred following secondary and tertiary infections. Lymphocytopaenia following primary CHMI is well-documented, (20) although further migration of cells from the periphery in repeat CHMI may reflect an immunological recall response upon secondary and tertiary infection, similar to our previous observations on antigen-specific B cells induced by subunit blood-stage vaccination prior to CHMI (21).

We also detected a derangement in liver transaminases following primary CHMI in approximately 25% of the participants (two in VAC063C and four in VAC063A and B), with the two in VAC063C of greatest severity and noticeably occurring at six days post-treatment initiation. This has not been previously observed in our centre, potentially due to lack of routine testing at the T+6 time point. Given that the abnormalities rapidly improved, these AEs would not have been detected at later follow-up visits, raising the possibility that frequency of liver function test (LFT) abnormalities post-primary CHMI may have been previously underestimated. LFT abnormalities are known to occur in both naturally acquired and experimental infection, and in blood-stage primary CHMI studies performed at QIMR, Australia, moderate and severe elevations of transaminase elevations enzymes have been reported, peaking at 4-12 days after antimalarial treatment, with ALT/AST ratio >1 and normal bilirubin in both *P. falciparum* and *P. vivax* infection models (22). Our findings are consistent with this pattern and given timing of onset and resolution, together with the observation that abnormal ALT levels normalise after treatment in severe malaria, suggest the transaminitis is likely to relate directly to CHMI, with inflammation/infection driving the ALT rather than the antimalarial drugs. However, since the transaminitis affected only a small subset of the primary CHMI cohort, individual host factors may also have contributed. Importantly, all ALT abnormalities were transient in nature, resolved spontaneously and were not associated with clinical symptoms or any derangement of synthetic liver function, so these findings do not raise clinical concern or preclude further testing using this repeat CHMI model, however, monitoring over similar time-points in future studies is warranted. Moreover, we also saw no derangement in LFTs after secondary or tertiary CHMI. We hypothesise that this could represent a degree of clinical immunity or immune “tolerance”, given transaminitis is a collateral marker of tissue damage. This is consistent with a modified CD4^+^ T cell response after a single drug-treated infection that has since been demonstrated in further work using these trial samples (23).

The blood-type of the donor of the red blood cells infected with 3D7 clone *P. falciparum* was Group O Rhesus negative, and the volume administered for CHMI is extremely small, equivalent to only a few microlitres of blood. As such, the risk of development of red cell alloantibodies following blood-stage CHMI was deemed to be very low. Nevertheless, as an exploratory measure, participants who were re-challenged in the VAC063A, B and C studies, and therefore re-exposed to the same inoculum, were retrospectively screened for IgG alloantibodies to red cells pre-CHMI and post their final follow-up. All samples tested were negative for red cell alloantibodies, consistent with single CHMI data from other groups using the same challenge inoculum (James McCarthy, personal communication). These data also suggest that an antibody response against the challenge inoculum itself was not a confounding factor in these re-challenge studies, which is also consistent with the very similar parasite growth kinetics observed in the majority of re-challengees.

Across the repeat CHMIs, all participants developed patent parasitaemia and reached diagnostic criteria, with the exception of the one participant who failed to reach diagnosis after secondary CHMI. By and large, our data suggest minimal to no acquisition of effective anti-parasitic blood-stage immunity occurs following one or two drug-treated and acute exposures to *P. falciparum*, even when using the homologous parasite clone. This is not inconsistent with observations from natural infection in the field, albeit here exposure to heterologous parasite strains will occur and blood-stage parasitaemias are likely to reach higher levels prior to treatment. This is also in stark contrast to the highly effective pre-erythrocytic immunity afforded by repeat exposure to sporozoite CHMI under drug cover (24, 25). We nonetheless were able to observe the priming and boosting of antibody responses to the well-studied merozoite antigens MSP1_19_ and AMA1, but not RH5. This hierarchy of immuno-dominance of *de novo* antibody induction is consistent with our previous observations in other primary CHMI trials (26, 27), and relatively short-lived responses boosted by each successive malaria exposure is consistent with observations from field studies spanning the malaria season (28). Notably our repeat CHMI data would strongly suggest these responses, as measured, remain well below a threshold that could contribute to reduced blood-stage parasite growth *in vivo*.

Nonetheless, we did note three participants of interest whose PMR varied across the repeat CHMIs. In one volunteer parasites grew consistently slowly, in another a modest reduction in PMR was seen upon secondary CHMI, and finally one volunteer showed a dramatic suppression of growth following secondary CHMI that was not repeated following tertiary CHMI. We could identify no demographic or obvious host factors to explain these observations, and these three volunteers had no history of prior malaria exposure. The latter participant’s only abnormality on blood tests was the development of a moderate anaemia by 28 days post-secondary CHMI, and their haemoglobin immediately preceding CHMI had been borderline low (111 g/dL). The relationship between iron status and malaria is complex (29) but it is unlikely that a short-term moderate anaemia would have caused such significant suppression of parasite growth. This is also supported by the fact that when a tertiary infection was administered the participant’s haemoglobin dropped again to similar levels but parasite growth conversely thrived. Also no systemic supplemental iron was known to be taken at the time of CHMI that could explain the increased growth rate. Instead, the highly anomalous PMR seen after secondary infection might be explained by one or more of the following: i) the induction and then waning of an effective immune response, or the induction of a highly specific immune response against a variant surface antigen on the infected red blood cell that was only expressed by the parasite during the secondary CHMI; ii) surreptitious or accidental self-treatment with a drug with an antimalarial effect; or iii) an operator error in the administration of the secondary challenge inoculum. The latter would seem unlikely, because even if inoculated with a single parasite, with 10-fold blood-stage growth every 48 hours, this would not lead to such prolonged Qpcr negativity in a non-immune subject. Regarding drugs, we could identify no such intervention, and performed an anti-malarial drug screen that was negative. Further studies would thus be needed to investigate if this was indeed a highly effective, but variant antigen-specific, immune response.

Overall, our data support the safety of repeat CHMI with blood-stage *P. falciparum* but suggest this approach will unlikely provide a model to study effective anti-parasitic immunity. On-going work, including with the recently established *P. vivax* blood-stage CHMI model in Oxford (30), will now focus on using the unique opportunities afforded by the CHMI system to interrogate mechanisms of immune tolerance and the acquisition of clinical immunity with the goal of informing next-generation approaches to protect against malaria disease.

## Methods

### Study Design and Participants

VAC063C was a non-blinded blood-stage *P. falciparum* controlled human malaria infection (CHMI) study to evaluate the safety and parasite growth dynamics of primary, secondary and tertiary blood-stage *P. falciparum* CHMI of healthy malaria-naïve UK adults. Healthy, malaria-naïve and non-pregnant adults aged 18-50 were invited to participate in the study. Volunteers were recruited and challenged at the Centre for Clinical Vaccinology and Tropical Medicine (CCVTM), part of the Oxford Vaccine Centre (OVC) at the University of Oxford. Eleven volunteers were enrolled in total. A full list of inclusion and exclusion criteria is reported in **Supplementary Methods**. Allocation to study group (Groups 1 and 2) was based on previous involvement and CHMI exposure in the VAC063 trial (ClinicalTrials.gov NCT02927145) (16), with new malaria-naïve volunteers comprising Group 3. The primary endpoint of the study was safety of repeat homologous CHMI (as measured by active and passive collection of clinical and laboratory AEs after each CHMI) and qPCR-derived parasite multiplication rate (PMR) was the primary endpoint for the assessment of parasite growth dynamics.

Data were pooled for analysis from participants who were enrolled in both VAC063C (November 2018) and in two previous CHMI studies under the preceding VAC063 protocol – VAC063A (November 2017) and VAC063B (March 2018). Briefly, the VAC063 protocol encompassed VAC063A and VAC063B, and was an open label, non-randomised Phase I/IIa clinical trial evaluating vaccine efficacy of the recombinant blood stage malaria protein RH5.1 in AS01_B_ adjuvant (16). The VAC063C trial was conducted to investigate the durability of any protective anti-parasite immunity measured in control (non-vaccinated) participants upon homologous re-challenge for the second or third time.

### Study Oversight

The VAC063C study was designed and conducted in the UK at the CCVTM, University of Oxford. The study was registered on ClinicalTrials.gov (NCT03906474) and received ethical approval from the UK National Health Service Research Ethics Services (South Central – Oxford A reference 18/SC/0521). All participants provided written informed consent and consent was verified before each CHMI. The study was conducted according to the principles of the current revision of the Declaration of Helsinki 2008 and in full conformity with the ICH guidelines for Good Clinical Practice (GCP). GCP compliance was independently and externally monitored by the Clinical Trials and Research Governance (CTRG) Team at the University of Oxford. Details of the previous VAC063 study approvals and oversight (from which primary and secondary CHMI data were used for a pooled analysis) are as previously described (16).

### Controlled Human Malaria Infection

The blood samples used as infectious inocula in this study were produced by Drs Gregor Lawrence, Allan Saul and colleagues at QIMR in Brisbane, Australia in 1994 and consist of aliquots of *P. falciparum* (clone 3D7) infected erythrocytes taken from a single donor (31). Blood was collected at the Australian Red Cross Blood Bank in an aseptic manner using standard blood bank equipment. The leukocytes were removed with a leukocytic filter. The thawing and washing of the cells reduced the amount of serum transferred with the red cells by a factor of 1000, compared to injecting the same volume of blood. The red cells were cryopreserved using a protocol from the American Association of Blood Banks Technical Manual that is normally employed for freezing blood from patients and donors with rare blood groups. The blood group of the donor was group O and Rhesus negative, Kell antigen negative. To date, CHMI of malaria-naïve individuals using this inoculum has always resulted in parasitaemia as detected by qPCR and/or microscopy (17, 19, 31, 32). Between 1994 and 2003 the cryopreserved samples to be used in this trial were stored in dedicated liquid nitrogen cylinders in a secure facility at QIMR. In 2003 the samples were transferred to Biotec Distribution Ltd., Bridgend, UK and then to Thermo Fisher Bishop’s Stortford, Hertfordshire, UK, in 2007 where they have been stored on behalf of the University of Oxford in temperature-monitored liquid nitrogen. Participants in VAC063C were each infected by direct intravenous inoculation of *P. falciparum* (clone 3D7) blood-stage parasites in the same way as for the VAC063A and B CHMI studies. The target inoculum dose was 1000 parasitised erythrocytes per participant. The inoculum was thawed and prepared under strict aseptic conditions as previously described (16) and volunteers received infected red cells in a total volume of 5 mL normal saline, followed by a saline flush. Subjects were observed for 1 h post-CHMI before discharge. The order in which volunteers from different groups were inoculated was interspersed in case of time effects on viability of the parasites. Following CHMI, blood samples were taken once on day one post-challenge (day C+1) and twice daily from day two (day C+2) for qPCR (target gene = 18S ribosomal RNA), to measure parasite density in real time.

Diagnosis of malaria was made on the basis of presence of symptoms in-keeping with malaria infection together with qPCR ≥5,000 parasites/mL (p/mL) or any available qPCR ≥10,000 p/mL, even if asymptomatic. Note in the VAC063A study, thick blood films were also evaluated at each time-point by experienced microscopists and diagnosis required volunteers to fulfil two out of three criteria: a positive thick blood film (one viable parasite in 200 fields) and/or qPCR ≥5,000 parasites/mL and/or symptoms consistent with malaria. In VAC063B and C microscopy was removed as a diagnostic tool to reduce the risk of participants being diagnosed prematurely, without any impact on participant safety. Participants were treated with a course of artemether/lumefantrine (Riamet) at diagnosis (n=9), or where contraindicated, with a course of atovaquone/proguanil (Malarone) (n=2). Half of the Riamet doses and all Malarone doses were directly observed by study investigators. After completion of treatment, follow-up visits were conducted at 6 days post-diagnosis/treatment (T+6), as well as at 28, 45 and 90 days post-CHMI (C+28, C+45 and C+90). This was similar though not identical to the follow-up schedule for VAC063A and B CHMIs where a T+6 visit was not included.

### Safety Analysis

Participants were reviewed once on day 1 post-challenge and twice-daily from day 2. At each visit they were asked a list of symptoms commonly associated with malaria infection (‘solicited’ symptoms including feverishness, malaise, fatigue, arthralgia, back pain, headache, myalgia, chills, rigors, sweats, headache, nausea, vomiting and diarrhoea). The severity of any reported symptom was then graded by the participant from 1 (mild) to 3 (severe), using the severity grading criteria shown in **Supplementary Material**. These were all recorded by the investigator as solicited AEs if they occurred during the 28 day period post-CHMI (or until completion of antimalarial treatment). Pyrexia was scored as follows: absent (_≤_ 37.5°C), mild (37.6 - 38.2°C), moderate (38.3 - 38.9°C) and severe (≥ 39°C) and participants were also asked to measure and record their temperature in a diary card if they experienced feverishness outside of their clinic visit. Blood samples for full blood count and biochemistry were obtained prior to CHMI (C-1) and then at C+6, diagnosis, T+6, C+45 and C+90. These were evaluated at Oxford University Hospitals NHS Trust providing 5-part differential white cell counts and quantification of electrolytes, urea, creatinine, bilirubin, alanine aminotransferase (ALT), alkaline phosphatase (ALP) and albumin. Blood was also tested serologically for evidence of Hepatitis B, Hepatitis C, HIV, EBV and CMV infection prior to CHMI. Blood tests were carried out at other time-points if clinically indicated.

All other (unsolicited) AEs were collected until day 90 post-CHMI and their likely causality in relation to CHMI or antimalarial drugs assessed and assigned a MedDRA code as described in the protocol. All AEs considered possibly, probably or definitely related to CHMI/antimalarial drugs were reported (**Table S2**), and all laboratory AEs at least possibly related to study interventions are also reported (**Table S3**).

### qPCR and PMR Modelling

Quantitative PCR was performed as previously reported for the VAC063A and B studies (16), and these data were used to model the PMR. The arithmetic mean of the three replicate qPCR results obtained for each individual at each timepoint was used for model-fitting. Negative individual replicates and data points which, based upon the mean of the three replicates, were negative or below the lower limit of quantification (20 parasites/mL), were handled as specified in the laboratory qPCR standard operating procedure. PMR was calculated using a linear model fitted to log_10_-transformed qPCR data (33). As previously, fitted lines were constrained to pass through the known starting parasitaemia, calculated from the results of a limiting-dilution-based assay of the number of viable parasites in the inoculum (34) and a weight-based estimate of each volunteer’s blood volume (70mL/kg) (35).

### Total IgG ELISAs

ELISAs to MSP1_19_, AMA1 and RH5 (all 3D7 sequence) were performed on serum samples using standardised methodology, all as previously described (16, 18).

### Red Cell Alloantibody Measurement

Serum samples were retrospectively screened for IgG alloantibodies to red cells (performed by Oxford University Hospitals NHS Foundation Trust using Capture-R® Ready-Screen® solid phase system) at C-1 (pre-CHMI) and C+90 (final post-CHMI follow-up).

### Antimalarial Drug Screen

Plasma taken both pre-CHMI and at day of diagnosis was sent for antimalarial compound testing at the Walter Reed Army Institute of Research (WRAIR) in the USA, using previously described mass spectrometry methodology. Samples were sent from two participants who demonstrated reduced parasite growth after repeat CHMI and from one participant who had the same growth rate across all three CHMIs for comparison. The antimalarial compounds tested included amodiaquine, artemisinin, atovaquone, chloroquine, clindamycin, doxycycline, lumefantrine, mefloquine, proguanil, pyrimethamine, quinine, and sulfadoxine.

### Statistical Analysis

Data were analysed using GraphPad Prism version 9.3.1 for Windows (GraphPad Software Inc., California, USA). Statistical tests used are reported in the Results text, and included two-tailed Kruskal-Wallis test with Dunn’s multiple comparison post-test, and log-rank analysis (Mantel Cox) of the Kaplan Meier curves. A value of *P* < 0.05 was considered significant.

## Data Availability

Requests for materials should be addressed to the corresponding author.

## Author Contributions

Conceived and performed the experiments: YT, JRB, CHM, TAR, ROP, MMH, BK, NJE, CMN, DMS, FAB, WN, RLR, MB, FR-L, PMF, DQ, KJE, IDP, PJS, SES, SJD, AMM

Analyzed the data: JS, YT, SES, JRB, NJE, SJD, AMM.

Project Management: JSC, FLN.

Wrote the paper: JS, YT, SES, SJD, AMM.

## Acknowledgments

The authors are grateful for the assistance of Natalie Lella and Michelle Kuskova for recruitment activities, Daniel Marshall Searson for database management, Julie Furze for laboratory assistance, and Richard Morter, Amy Flaxman and Duncan Bellamy for qPCR support (Jenner Institute, University of Oxford); Amy Duckett and Carly Banner for arranging contracts (University of Oxford); James McCarthy for providing the 3D7 inoculum (QIMR, Australia); Amy Noe (Leidos Life Sciences) and Lorraine Soisson (U.S. Agency for International Development, USAID) for useful discussions; William Dennis and Chau Vuong (WRAIR, USA) for assisting with the antimalarial drug testing; and all the study volunteers. The VAC063 clinical trial was funded by the Office of Infectious Diseases, Bureau for Global Health, USAID, under the terms of Malaria Vaccine Development Program (MVDP) contract AID-OAA-C-15-00071, for which Leidos, Inc. was the prime contractor. The opinions expressed here are those of the authors and do not necessarily reflect the views of the USAID. This work was also supported in part by the National Institute for Health Research (NIHR) Oxford Biomedical Research Centre (BRC). The views expressed are those of the authors and not necessarily those of the NIHR or the Department of Health and Social Care. TAR held a Wellcome Trust Research Training Fellowship [108734/Z/15/Z] and CMN held a Wellcome Trust Sir Henry Wellcome Postdoctoral Fellowship [209200/Z/17/Z]. SJD is a Jenner Investigator and held a Wellcome Trust Senior Fellowship [106917/Z/15/Z].

## Data and Materials Availability

Requests for materials should be addressed to the corresponding author.

