## Supplementary Materials for "Repeat controlled human malaria infection of healthy UK adults with blood-stage *Plasmodium falciparum*: safety and parasite growth dynamics"

- Supplementary Methods
  - Inclusion and exclusion criteria for the VAC063C study
  - Severity Grading of AEs
  - Causality Assessment
  - Severity grading criteria for clinically significant laboratory abnormalities
- Supplementary Figures and Tables
  - Table S1. Demographics for study participants
  - Table S2. Unsolicited AEs considered possibly, probably or definitely related to study interventions (CHMI or antimalarials)
  - Table S3. Laboratory AEs considered possibly, probably or definitely related to study interventions
  - Table S4. Raw qPCR data for VAC063C study
  - Figure S1. AEs considered possibly, probably or definitely related to antimalarial therapy for VAC063A, B and C
  - Figure S2. Alanine aminotransferase (ALT) over time for VAC063 A and B

### Supplementary Methods

#### Inclusion and Exclusion Criteria

A medical history and physical examination were conducted at the screening visit, as well as baseline blood tests including a full blood count; urea and electrolytes; liver function tests; and hepatitis B virus (HBV), hepatitis C virus (HCV) human immunodeficiency virus (HIV). Epstein-Barr virus (EBV) and Cytomegalovirus (CMV) serology. Dipstick urinalysis for all volunteers and pregnancy testing for all female volunteers were conducted at screening. Pregnancy testing (serum Beta Human Chorionic Gonadotropin, Beta hCG), was also carried out prior to CHMI. A full list of inclusion and exclusion criteria is shown below:

##### Inclusion Criteria

This study will be conducted in healthy adults, who meet the following inclusion and exclusion criteria:

**Inclusion Criteria**

The volunteer must satisfy all the following criteria to be eligible for the study:

- Healthy adults aged 18 to 50 years.
- Able and willing (in the Investigator’s opinion) to comply with all study requirements.
- Willing to allow the Investigators to discuss the volunteer’s medical history with their General Practitioner (GP).
- For females only, willingness to practice continuous effective contraception (see below) during the study and a negative pregnancy test on the day(s) of screening and on the day prior to blood-stage CHMI, and prior to the start of antimalarial treatment.
- Provide written informed consent.
- Agreement to permanently refrain from blood donation, as per current UK Blood Transfusion and Tissue Transplantation Services guidelines
- Reachable (24 hours a day) by mobile phone during the period between CHMI and completion of antimalarial treatment
- Willingness to take a curative anti-malarial regimen following CHMI.
- Answer all questions on the informed consent questionnaire correctly.
- For Groups 1-2: completion of primary or secondary challenge in the VAC063 trial, curative anti-malarials and follow-up (up until at least the C+28 visit).

**Exclusion Criteria**

The volunteer may not enter the study if any of the following apply:

- Participation in another research study involving receipt of an investigational product in the 30 days preceding enrolment, or planned use during the study period.
- Prior receipt of an investigational vaccine likely to impact on interpretation of the trial data, as assessed by the Investigator.
- Administration of immunoglobulins and/or any blood products at any time in the past.
- Any confirmed or suspected immunosuppressive or immunodeficient state, including HIV infection; asplenia; recurrent, severe infections and chronic (more than 14 days) immunosuppressant medication within the past 6 months (inhaled and topical steroids are allowed).
- Administration of long-acting immune-modifying drugs at any time during the study period (e.g. infliximab).
- History of malaria chemoprophylaxis within 30 days prior to CHMI.
- Use of systemic antibiotics with known antimalarial activity within 30 days of CHMI (e.g. trimethoprim-sulfamethoxazole, doxycycline, tetracycline, clindamycin, erythromycin, fluoroquinolones and azithromycin).
- History of allergic disease or reactions likely to be exacerbated by malaria infection.
- Pregnancy, lactation or willingness/intention to become pregnant during the study.
- History of cancer (except basal cell carcinoma of the skin and cervical carcinoma in situ).
- History of serious psychiatric condition likely to affect participation in the study.
- Any other serious chronic illness requiring hospital specialist supervision.
- Suspected or known current alcohol abuse as defined by an alcohol intake of greater than 25 units every week.
- Suspected or known injecting drug abuse in the 5 years preceding enrolment.
- Seropositive for hepatitis B surface antigen (HBsAg) at screening.
- Seropositive for HIV virus (antibodies to HIV) at screening
- Seropositive for hepatitis C virus (antibodies to HCV) at screening (***unless*** has taken part in a prior hepatitis C vaccine study with confirmed negative HCV antibodies prior to participation in that study, and negative HCV RNA PCR at screening for this study).
- History of clinical malaria (any species - NOT applicable to prior challenge in VAC063 study for Groups 1 and 2).
- Travel to a malaria endemic region during the study period or within the previous six months.
- Any clinically significant abnormal finding on screening biochemistry or haematology blood tests or urinalysis.
- Any other significant disease, disorder or finding which may significantly increase the risk to the volunteer because of participation in the study, affect the ability of the volunteer to participate in the study or impair interpretation of the study data.
- Inability of the study team to contact the volunteer’s GP to confirm medical history to allow Investigator to assess safety to participate.
- History of sickle cell anaemia, sickle cell trait, thalassaemia or thalassaemia trait or any haematological condition that could affect susceptibility to malaria infection.
- Laboratory evidence of G6PD deficiency at screening.
- Laboratory evidence of haemoglobinopathy at screening.
- Use of medications known to cause prolongation of the QT interval ***and*** existing contraindication to the use of Malarone.
- Use of medications known to have a potentially clinically significant interaction with Riamet ***and*** Malarone.
- Contraindications to the use of **both** Riamet ***and*** Malarone.
- Any clinical condition known to prolong the QT interval.
- Family history of congenital QT prolongation or sudden death.
- Positive family history in **both** 1st and 2nd degree relatives < 50 years old for cardiac disease.
- History of cardiac arrhythmia, including clinically relevant bradycardia.
- Volunteer unable to be closely followed for social, geographic or psychological reasons.

**Effective contraception for female volunteers**

Female volunteers are required to use an effective form of contraception during the course of the study.

Acceptable forms of contraception for female volunteers include:

- Established use of oral, injected or implanted hormonal methods of contraception.
- Placement of an intrauterine device (IUD) or intrauterine system (IUS).
- Barrier methods of contraception (condom or occlusive cap with spermicide).
- Male sterilisation, if the vasectomised partner is the sole partner for the subject.
- True abstinence, when this is in line with the preferred and usual lifestyle of the subject (periodic abstinence and withdrawal are not acceptable methods of contraception).

N.B.: Women using hormonal contraceptives during CHMI and treated with the antimalarial Riamet will be advised to also use a barrier method of contraception whilst on Riamet treatment, and until the start of the next menstruation after treatment.

**Severity Grading of AEs**

Participants graded all AEs as mild, moderate or severe:

- **GRADE 0:** None.
- **GRADE 1:** Transient or mild discomfort (< 48 h); no medical intervention/therapy required.
- **GRADE 2:** Mild to moderate limitation in activity – some assistance may be needed; no or minimal medical intervention/therapy required.
- **GRADE 3:** Marked limitation in activity, some assistance usually required; medical intervention/therapy required; hospitalization possible.

Adverse event data also included the results of the haematology (full blood count) and biochemistry (liver function tests, urea and electrolytes).

**Causality Assessment**

For each unsolicited AE, an assessment of the relationship of the AE to the study intervention(s) was undertaken. Alternative causes of the AE, such as the natural history of pre-existing medical conditions, concomitant therapy, other risk factors and the temporal relationship of the event to CHMI/ antimalarials were considered. The likely causality of all unsolicited AEs was assessed as per the criteria below:

- **No Relationship:** No temporal relationship to study intervention ***and*** alternate aetiology (clinical state, environmental or other interventions); ***and*** does not follow known pattern of response to study intervention.
- **Unlikely:** Unlikely temporal relationship to study intervention ***and*** alternate aetiology likely (clinical state, environmental or other interventions) ***and*** does not follow known typical or plausible pattern of response to study intervention.
- **Possible:** Reasonable temporal relationship to study intervention; ***or*** event not readily produced by clinical state, environmental or other interventions; ***or*** similar pattern of response to that seen with other similar interventions.
- **Probable:** Reasonable temporal relationship to study intervention; ***and*** event not readily produced by clinical state, environment, or other interventions ***or*** known pattern of response seen with other similar interventions.
- **Definite:** Reasonable temporal relationship to study intervention; ***and*** event not readily produced by clinical state, environment, or other interventions; ***and*** known pattern of response seen with other similar interventions.

All unsolicited AEs that were assessed as being possibly, probably or definitely related to either CHMI or antimalarials are shown in **Table S2**.

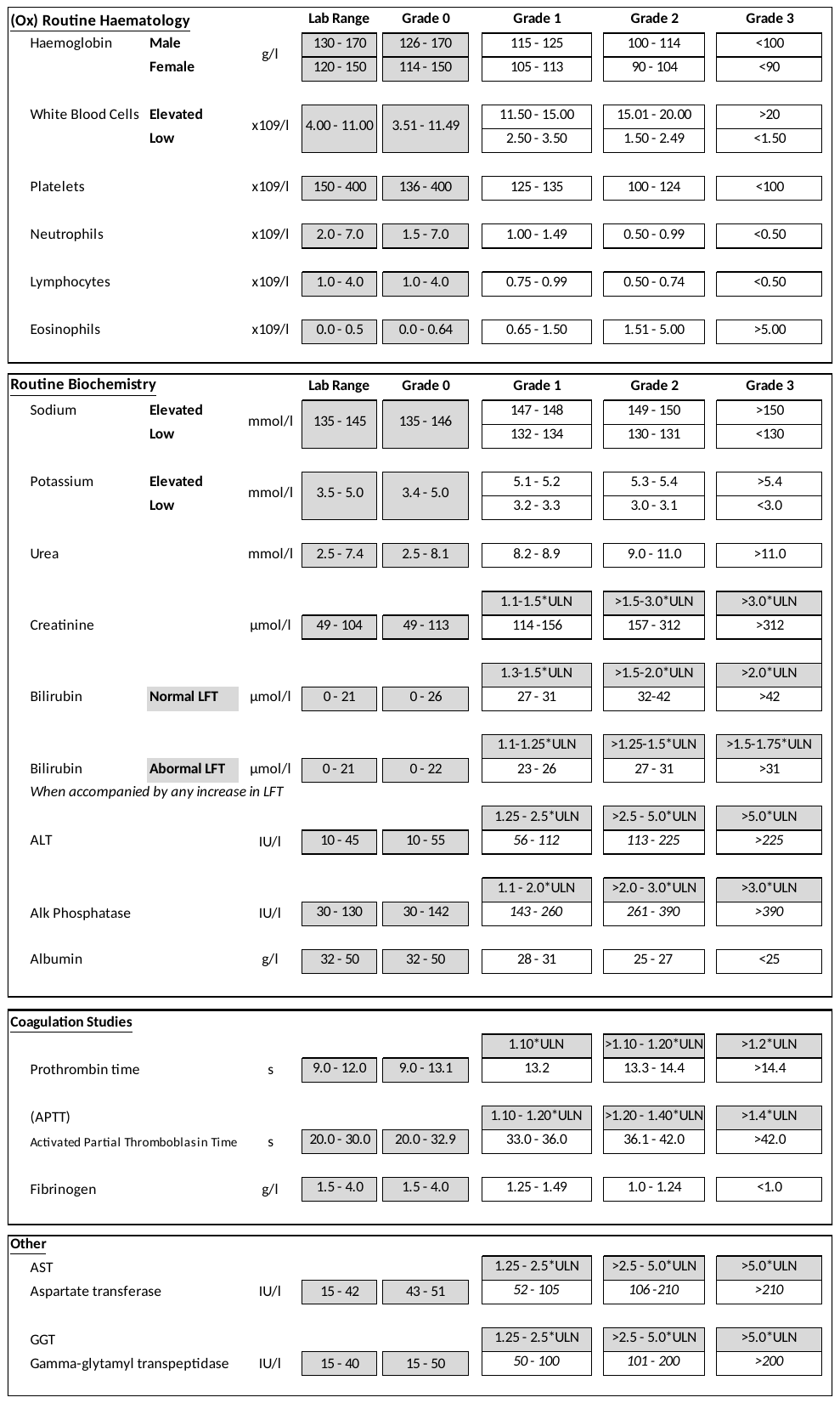

**Severity grading criteria for clinically significant laboratory abnormalities;** adapted from FDA guidelines using Oxford University Hospitals NHS Foundation Trust laboratory reference ranges.

### Supplementary Tables and Figures

#### Table S1:  Demographic information for study participants.

Information is shown for VAC063C participants plus pooled data across VAC063A, B and C CHMIs.

|  |  | **VAC063C** | | | **All participants** | | |
| --- | --- | --- | --- | --- | --- | --- | --- |
|  |  | **Primary CHMI** | **Secondary CHMI** | **Tertiary CHMI** | **Primary CHMI** | **Secondary CHMI** | **Tertiary CHMI** |
| **No. of participants** | | 3 | 2 | 6 | 24 | 10 | 6 |
| **Sex: no. female** | | 2 (67%) | 2 (100%) | 2 (33%) | 12 (50%) | 4 (40%) | 2 (33%) |
| **Age (years)** | **Median** | 25 | 26 | 29 | 26.5 | 23.5 | 29 |
|  | **Range** | 23-50 | 23-29 | 23-34 | 20-50 | 21-34 | 23-34 |
| **Body mass index (kg/m^2^)** | **Median** | 29.8 | 19.3 | 24.6 | 24.8 | 25.2 | 24.6 |
|  | **Range** | 24.4-31.5 | 19.1-19.5 | 19.0-33.0 | 18.7-39.0 | 19.1-33.4 | 19.0-33.0 |
| **Ethnicity** | **White British** | 2 (67%) | 2 (100%) | 4 (67%) | 14 (58%) | 8 (80%) | 4 (67%) |
|  | **White other** | 0 (0%) | 0 (0%) | 1 (17%) | 5 (21%) | 1 (10%) | 1 (17%) |
|  | **Asian** | 0 (0%) | 0 (0%) | 1 (17%) | 3 (13%) | 1 (10%) | 1 (17%) |
|  | **Other** | 1 (33%) | 0 (0%) | 0 (0%) | 2 (8%) | 0 (0%) | 0 (0%) |
| **Smoking** | | Unknown | 1 (50%) | 1 (17%) | 2/21 (9.5%) | 2 (20%) | 1 (17%) |
| **Alcohol excess** | | Unknown | 0 (0%) | 0 (0%) | 0/21 (0%) | 0 (0%) | 0 (0%) |

#### Table S2: Unsolicited AEs.

Unsolicited AEs deemed at least possibly (possibly, probably or definitely) related to study interventions in all volunteers in VAC063A, B and C are shown, with MedDRA coding and maximum severity reported. (**A**) Unsolicited AEs deemed at least possibly related to CHMI. (**B**) Unsolicited AEs deemed at least possibly related to study drugs. Groups 3, 6 and 9 underwent primary CHMI; Groups 2 and 8 underwent secondary CHMI; Group 1 underwent tertiary CHMI.

**Table S2A**

| **MedDRA System Organ Class** | **MedDRA Higher Level Term** | **MedDRA Preferred Term** | **Group** | **Onset (days post-CHMI)** | **Duration (days)** | **Maximum severity** |
| --- | --- | --- | --- | --- | --- | --- |
| **Nervous system disorders** | Headaches NEC | Headache | 1 | 14 | 3 | 1 |
|  |  |  | 9 | 13 | 3 | 3 |
| **General disorders and administration site conditions** | Asthenic conditions | Malaise | 6 | 12 | 3 | 2 |
|  |  | Pyrexia | 6 | 11 | 1 | 2 |
|  | General signs and symptoms NEC | Exercise tolerance decreased | 1 | 13 | 3 | 1 |
|  | Pain and discomfort NEC | Chest pain | 9 | 8 | 2 | 1 |
| **Gastrointestinal disorders** | Gastrointestinal and abdominal pains (excl oral and throat) | Abdominal pain | 2 | 7 | 1 | 2 |
|  |  |  | 2 | 8 | 0 | 2 |
|  |  |  | 6 | 3 | 0 | 2 |
|  |  |  | 6 | 8 | 8 | 2 |
|  |  |  | 6 | 9 | 4 | 1 |
|  |  | Abdominal pain upper | 8 | 13 | 2 | 1 |
|  | Dyspeptic signs and symptoms | Dyspepsia | 8 | 11 | 3 | 2 |
| **Cardiac disorders** | Cardiac signs and symptoms NEC | Dizziness | 2 | 10 | 1 | 3 |
|  |  |  | 2 | 13 | 2 | 2 |
|  |  |  | 2 | 9 | 1 | 1 |
|  |  |  | 8 | 2 | 1 | 2 |
| **Metabolism and nutrition disorders** | Appetite disorders | Decreased appetite | 3 | 8 | 33 | 1 |
|  |  |  | 9 | 11 | 5 | 2 |
|  | Iron deficiencies | Iron deficiency | 6 | 17 | 12 | 2 |
|  | Total fluid volume decreased | Dehydration | 9 | 9 | 0 | 1 |
| **Psychiatric disorders** | Dyssomnias | Somnolence |  | 7 | 2 | 1 |
| **Skin and subcutaneous tissue disorders** | Dermal and epidermal conditions NEC | Sensitive skin | 9 | 9 | 1 | 1 |
|  |  |  |  | 7 | 6 | 1 |

**Table S2B**

| **MedDRA System Organ Class** | **MedDRA Higher Level Term** | **MedDRA Preferred Term** | **Group** | **Onset (days post-CHMI)** | **Duration (days)** | **Maximum severity** |
| --- | --- | --- | --- | --- | --- | --- |
| **General disorders and administration site conditions** | General signs and symptoms NEC | Exercise tolerance decreased | 1 | 13 | 3 | 1 |
| **Metabolism and nutrition disorders** | Appetite disorders | Decreased appetite | 3 | 8 | 33 | 1 |
| **Nervous system disorders** | Paraesthesias and dysaesthesias | Hypoaesthesia | 3 | 11 | 0 | 1 |

#### Table S3:  Laboratory AEs.

All adverse events at least possibly related to study procedures for (**A**) primary CHMI, (**B**) secondary CHMI, and (**C**) tertiary CHMI. Laboratory abnormalities which were unlikely or not related to study procedures are not included (cf. **Figure 4A** which presents all abnormal haemoglobin, platelet and lymphocyte count results regardless of relatedness to CHMI). Of note, a moderate anaemia was seen post-CHMI in two individuals – one at C+28 post-primary CHMI and one at C+28 following secondary CHMI. The first was transient and had self-resolved by the next visit (C+90). The second improved over the period of study monitoring but was still ongoing at the final secondary CHMI study visit (haemoglobin 104 g/dL – Grade 2) where it was noted that the participant had a vegan diet and their pre-enrolment haemoglobin had also been borderline prior to secondary CHMI (111 g/dl). The participant was referred to their GP and subsequently started on iron supplements. Their haemoglobin recovered with short-term supplementary iron and they were then re-enrolled into VAC063C for a tertiary CHMI, with a pre-enrolment haemoglobin of 114 g/dL. By day of diagnosis their haemoglobin had fallen to 104 g/dL (Grade 2) again, and although it once again improved it remained at Grade 1 at the final study visit.

**Table S3A**

| **Laboratory AE** | **Percent participants with Lab AE (n=24)** | **Group** | **Timepoint of onset** | **Max. severity** | **Relatedness to study intervention** | **Resolved by C+90** |
| --- | --- | --- | --- | --- | --- | --- |
| Anaemia | 21% | 9 | C+28 | Grade 1 | Possible | Y |
|  |  | 6 | DoD | Grade 1 | Possible | Y |
|  |  | 6 | C+28 | Grade 1 | Possible | Y |
|  |  | 6 | C+28 | Grade 2 | Possible | Y |
|  |  | 6 | C+6 | Grade 1 | Possible | Y |
| Thrombocytopaenia | 8% | 3 | T+6 | Grade 2 | Probable | Y |
|  |  | 9 | DoD | Grade 2 | Probable | Y |
| Leucopaenia | 4% | 6 | C+28 | Grade 1 | Possible | Y |
| Leucocytosis | 8% | 6 | C+28 | Grade 1 | Possible | Y |
|  |  | 6 | DoD | Grade 2 | Possible | Y |
| Neutropaenia | 4% | 3 | T+6 | Grade 1 | Possible | Y |
| Lymphocytopaenia | 29% | 9 | DoD | Grade 3 | Probable | Y |
|  |  | 9 | DoD | Grade 3 | Probable | Y |
|  |  | 9 | DoD | Grade 3 | Probable | Y |
|  |  | 6 | DoD | Grade 1 | Probable | Y |
|  |  | 3 | DoD | Grade 2 | Probable | Y |
|  |  | 3 | DoD | Grade 2 | Probable | Y |
|  |  | 6 | DoD | Grade 2 | Probable | Y |
| Eosinophilia | 8% | 6 | C+6 | Grade 1 | Possible | Y |
|  |  | 6 | C+6 | Grade 1 | Possible | Y |
| Hypokalaemia | 21% | 9 | C+6 | Grade 1 | Possible | Y |
|  |  | 9 | DoD | Grade 1 | Possible | Y |
|  |  | 6 | DoD | Grade 3 | Possible | Y |
|  |  | 6 | DoD | Grade 1 | Possible | N |
|  |  | 6 | DoD | Grade 1 | Possible | Y |
| Elevated alanine aminotransaminase (ALT) | 25% | 3 | T+6 | Grade 3 | Probable | Y |
|  |  | 3 | T+6 | Grade 2 | Probable | Y |
|  |  | 6 | T+1  (additional visit) | Grade 1 | Probable | Y |
|  |  | 6 | C+28 | Grade 2 | Probable | Y |
|  |  | 9 | C+28 | Grade 1 | Possible | Y |
|  |  | 9 | T+7 (additional visit) | Grade 1 | Possible | Y |
| Elevated aspartate aminotransaminase (AST) | 8% | 3 | T+8 (additional visit) | Grade 2 | Probable | Y |
|  |  | 3 | T+8 (additional visit) | Grade 1 | Probable | Y |
| Elevated gamma-glutamyltransferase (GGT) | 8% | 3 | T+8 (additional visit) | Grade 3 | Probable | Y |
|  |  | 3 | T+8 (additional visit) | Grade 2 | Probable | Y |
| Elevated urea | 4% | 6 | C+28 | Grade 1 | Possible | Y |

**Table S3B**

| **Laboratory AE** | **Percent participants with Lab AE (n=10)** | **Group** | **Timepoint of onset** | **Max. severity** | **Relatedness to study intervention** | **Resolved by C+90** |
| --- | --- | --- | --- | --- | --- | --- |
| Anaemia | 20% | 2 | T+6 | Grade 1 | Possible | Y |
|  |  | 8 | C+28 | Grade 2 | Possible | N |
| Leucopaenia | 10% | 8 | DoD | Grade 1 | Probable | Y |
| Lymphocytopaenia | 60% | 2 | DoD | Grade 3 | Probable | Y |
|  |  | 2 | DoD | Grade 2 | Probable | Y |
|  |  | 8 | DoD | Grade 3 | Probable | Y |
|  |  | 8 | DoD | Grade 2 | Probable | Y |
|  |  | 8 | DoD | Grade 1 | Probable | Y |
|  |  | 8 | DoD | Grade 3 | Probable | Y |
| Low albumin | 10% | 8 | DoD | Grade 1 | Possible | Y |
| Elevated urea | 10% | 8 | C+6 | Grade 1 | Possible | Y |

**Table S3C**

| **Laboratory AE** | **Percent participants with Lab AE (n=6)** | **Group** | **Timepoint of onset** | **Max. severity** | **Relatedness to study intervention** | **Resolved by C+90** |
| --- | --- | --- | --- | --- | --- | --- |
| Anaemia | 17% | 1 | DoD | Grade 2 | Probable | N |
| Lymphocytopaenia | 67% | 1 | DoD | Grade 3 | Probable | Y |
|  |  | 1 | DoD | Grade 2 | Probable | Y |
|  |  | 1 | DoD | Grade 2 | Probable | Y |
|  |  | 1 | DoD | Grade 1 | Probable | Y |

#### Table S4: Raw qPCR data (parasites/mL) for VAC063C.

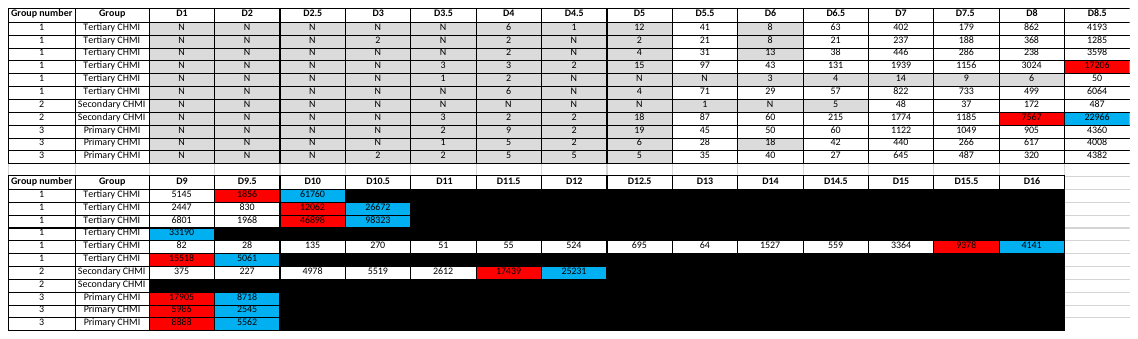
Top row represents day of follow-up visit post blood-stage CHMI. Data highlighted in red represent qPCR measurement on day of diagnosis (DoD) for a particular individual. N = PCR negative for all three triplicate readings in the assay. Squares highlighted in grey indicate negative or <20 p/mL below minimum positive reporting criteria. Squares highlighted in blue indicate an additional post-diagnosis sample. Note DoD indicated as diagnosis timepoint, but some individuals had their DoD sample taken at the next clinic and hence why there is an additional post-diagnosis (but pre-treatment) sample available.

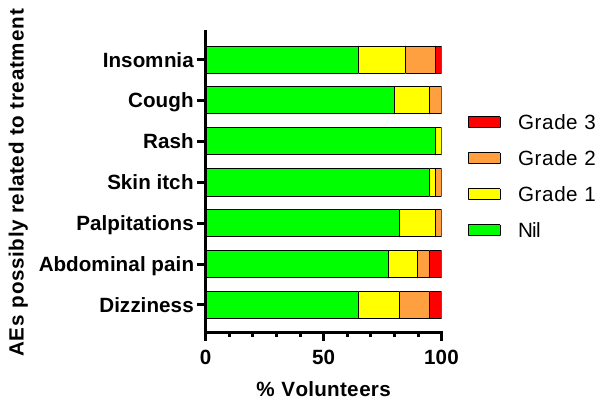

#### Figure S1: Safety of malaria treatment.

Frequency and severity of solicited AEs possibly related to malaria treatment across the groups in VAC063A, B and C (n=40).

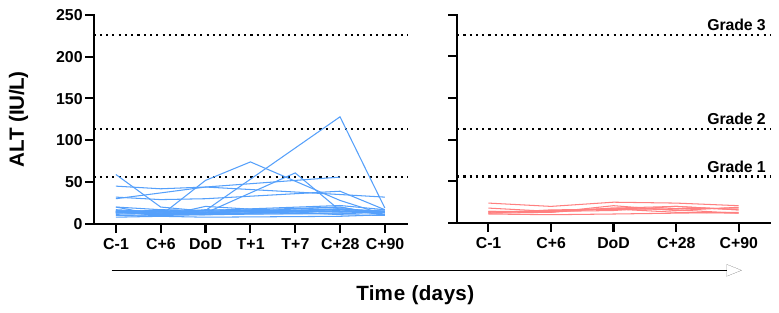

#### Figure S2: Alanine aminotransferase (ALT) over time for VAC063A and B.

ALT over time for each participant for primary (n=21) and secondary (n=8) CHMI. Dashed line shows the local cut off for Grade 1, 2 and 3 abnormalities. Time is number of days post-CHMI, except for day of diagnosis (DoD) which varies by participant. Unlike in VAC063C (**Figure 4B**), VAC063A and B did not include a day 6 post-treatment timepoint, but additional bleeds taken for two participants at day 1 (T+1) and day 7 (T+7) post-treatment are shown here. *P* value as calculated by Kruskal-Wallis test for maximum ALT result per participant showed no significant difference between primary, secondary and tertiary CHMI when all participants across VAC063A, B and C were included (*P* = 0.20).
